# Comparing computable structured phenotype- versus large language model-identification of opioid use disorder using electronic health record data

**DOI:** 10.64898/2025.12.17.25342510

**Authors:** Melanie Molina, Cynthia Fenton, Kathy T. LeSaint, Samuel D. Pimentel, Michael A. Kohn, Aaron E. Kornblith

## Abstract

**Study Objective:** To compare a computable structured opioid use disorder (OUD) phenotype currently used to trigger emergency department (ED) clinical decision support (CDS) with a large language model (LLM) for OUD identification, using expert physician review as the reference standard.

**Methods:** We conducted a retrospective study of randomly sampled adult ED encounters (January 1, 2023-October 17, 2024) at a single academic health system. Encounters were stratified by structured phenotype status and weighted to reflect population prevalence. The structured phenotype, implemented operationally to activate CDS, incorporated diagnosis codes, medications for OUD, urine toxicology results, addiction consultations, and keyword recognition. An LLM (ChatGPT 4.1) analyzed ED notes from the index visit using a zero-shot prompt. Two board-certified emergency physicians independently determined OUD status by full chart review; discrepancies were adjudicated by a third reviewer. We calculated weighted sensitivity, specificity, positive predictive value (PPV), and negative predictive value (NPV).

**Results:** Among 302 encounters, weighted OUD prevalence was 5.6% (95% CI 4.0-7.0%). The structured phenotype demonstrated sensitivity 0.84 (95% CI 0.42-0.97) and specificity 0.964 (95% CI 0.96-0.97) (PPV 0.58; NPV 0.99). The LLM demonstrated sensitivity 0.81 (95% CI 0.70-0.88) and specificity 0.996 (95% CI 0.993-0.998) (PPV 0.92; NPV 0.99). Specificity was significantly higher for the LLM (p<0.0001).

**Conclusion:** Both approaches demonstrated strong diagnostic performance. Although the structured phenotype showed slightly higher sensitivity, the LLM achieved higher specificity and PPV, suggesting potential to reduce false-positive alerts in ED workflows. Prospective validation in larger, representative populations is needed to guide clinical implementation.

## INTRODUCTION

### Background

Opioid use disorder (OUD) affects over 2.5 million Americans^1^ and contributed to more than 80,000 overdose deaths in 2022,^2^ constituting a critical public health crisis. The emergency department (ED) serves as a vital access point for patients with OUD,^3^ whether presenting with OUD-related or unrelated conditions. This creates an opportunity to connect patients to treatment and support.

### Importance

Screening for OUD in the ED is often limited due to competing clinical priorities, time constraints, and resource limitations.^4^ Failure to identify OUD during the ED visit represents a missed opportunity to initiate treatment and link patients to care.^5^ To address this challenge, prior work has developed computable structured OUD phenotypes within the electronic health record (EHR) to support screening and clinical decision-making.^6^ These approaches, validated across multiple health systems,^6^ show promise by potentially improving workflow efficiency and bypassing the need for manual screening by busy clinicians. In some settings, such phenotypes have been operationalized to activate real-time alerts or care coordination pathways, demonstrating feasibility for integration into routine ED workflows.

Nevertheless, computable structured OUD phenotypes may function as blunt instruments, missing clinical nuance in narrative documentation and potentially generating excess false positives in screening applications. Recent advances in artificial intelligence (AI) offer additional promise for improving OUD identification. Although AI approaches show promise for identifying OUD,^7^ their performance in ED settings, particularly generative large language models (LLM) used without task-specific training, remains largely untested. Such LLMs may capture clinical nuance in narrative documentation that rule-based screening tools miss.

As EHR vendors begin integrating LLM-based tools into ED clinical workflows, there is increasing opportunity to evaluate whether they can support scalable OUD identification. However, integration of LLMs into clinical care requires understanding their performance and limitations.

### Goals of This Investigation

We sought to compare a computable structured OUD phenotype against an LLM for OUD identification, using expert human determination as a reference standard.

## METHODS

### Study Design and Selection of Participants

We performed a retrospective analysis of EHR data extracted from the University of California, San Francisco (UCSF) Medical Center, with IRB approval. We began with all ED encounters for patients ≥18 years between January 1, 2023 and October 17, 2024. Given OUD is relatively rare in our patient population, we used stratified sampling to generate an enriched sample of encounters for application of the reference standard. We first stratified encounters by a computable structured OUD phenotype^8^ currently implemented within our health system (described below). Within each of the two strata, we randomly selected one encounter per unique patient, eliminating duplicates. We then performed random sampling to obtain 302 patient-encounters: 201 meeting the structured OUD phenotype and 101 not meeting the phenotype. The sample size was selected to ensure at least 100 with OUD by the reference standard. Each included ED encounter was required to have associated ED notes available for LLM analysis.

### Measurements

#### Computable Structured Phenotype

To identify vulnerable patients who could benefit from social work and care coordination, we employed a broad approach to capture patients with clinical evidence of OUD even without a formal diagnosis. We adapted the structured OUD phenotype originally developed and validated by Chartash et al.^6^ for our local implementation as part of a clinical decision support (CDS) tool designed to identify high-risk OUD patients with unmet social needs. The original phenotype was enhanced to improve sensitivity and accommodate our institutional EHR structure, as OUD documentation is often incomplete due to stigma and variable clinical practices.

Our site-adapted structured OUD phenotype^8^ triggers CDS if any of the following are true: ≥1 ICD-10 codes indicating OUD, withdrawal, or overdose in encounter (discharge and/or admission) diagnoses, problem lists, or past medical history; active prescriptions for buprenorphine or methadone preceding the index ED visit encounter; ≥1 positive urine toxicology for methadone, fentanyl, or heroin; ≥1 addiction care team consultation notes mentioning opioid-related terms; and text recognition of specific terms (“OUD,” “opioid use disorder,” “COWS,” “heroin,” “fentanyl”) in ED clinical notes, including reason for visit free text.

#### Large Language Model

We used OpenAI ChatGPT 4.1 with all notes associated with the ED encounter (excluding student and procedure notes) concatenated into the prompt without additional labels other than note titles. The prompt asked the model to determine whether the patient had OUD based on the clinical documentation and was iteratively defined based on ten test cases (**Supplementary Material Appendix A**).

#### Human Reference Standard

Two board-certified emergency medicine attending physicians, one with additional certification in addiction medicine and toxicology (KL) and the other in clinical informatics (MM), independently and manually reviewed each patient’s entire medical record, including Care Everywhere documentation, blinded to the results of both the structure phenotype and the LLM evaluation, to determine OUD status. They were deliberately given full chart access to reflect real-world clinical practice. A positive OUD status was defined as any of the following: 1) any documented history of OUD, 2) any history of opioid withdrawal, 3) any history of opioid overdose via intentional use of an opioid, 4) >1 episode of intentional opioid use. Opioid dependence was excluded. Discrepancies were adjudicated by a third attending physician (AK).

### Analysis

We used descriptive statistics to characterize ED encounters by OUD status and assessed interrater agreement using Cohen’s κ with 95% confidence intervals (CI). Stratified random sampling with the two strata defined by an index test is test result-based sampling.^9,10^ For the structured phenotype, the naively calculated PPV and NPV are unbiased, but estimating sensitivity and specificity requires weights equal to the inverse sampling fraction. For LLM, all metrics, PPV, NPV, sensitivity, and specificity require inverse sampling weights. We used Stata’s survey procedures with the finite population correction to calculate the weighted estimates with 95% confidence intervals. Differences in sensitivity and specificity between tests were assessed with survey-weighted paired difference-in-proportions Wald tests.

Analyses were conducted in R version 4.4.2 and Stata version 9.5. We performed qualitative analysis of discordant cases using rapid content analysis.

## RESULTS

From January 1, 2023 and October 17, 2024, there were 70880 encounters by adults > 18 years, 5701 (8.0%) structured phenotype-positive. Of these, 201 (sampling fraction 0.0353) were randomly selected for application of the reference standard. Of the remaining 65179 encounters that were structured phenotype-negative, 101 were randomly selected (sampling fraction 0.0015). In this sample of 302 encounters, 67 (22%) had multiple ED note types. Patient demographic characteristics are shown in **Supplementary Material Appendix B**.

There was strong interrater reliability (κ = 0.92, 95% CI 0.88-0.97) between the two physicians’ classifications with 10 encounters requiring adjudication. After consensus, 117 (38.7%) encounters were classified as having OUD and 185 (61.3%) as not having OUD. The test characteristics of the structured phenotype and LLM compared to the reference standard are shown in **Table 1**.

**Table 1.**
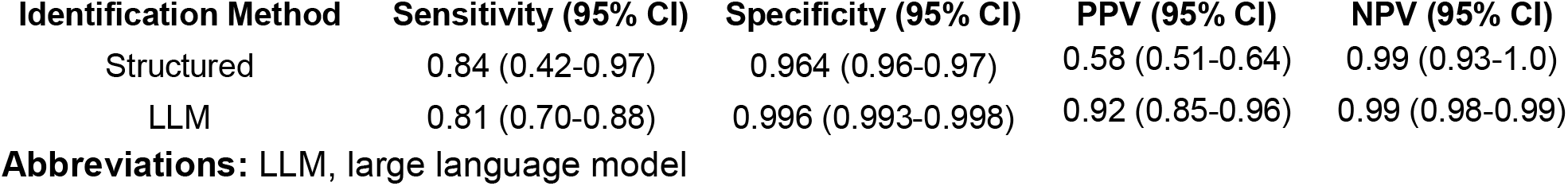
Test Characteristics of Computable Structured Phenotype versus LLM.

Compared to the LLM, the computable structured phenotype demonstrated comparable sensitivity, 0.84 vs. 0.81 (p = 0.859), and worse specificity, 0.964 vs. 0.996 (p < 0.0001). The estimated population prevalence of OUD was 0.056 (95% CI 0.040 to 0.07).

Qualitative analysis of discordant cases (**Table 2**) revealed distinct error patterns for each approach. Computable structured phenotype false positives primarily resulted from single positive fentanyl urine toxicology results and chronic pain patients on methadone or buprenorphine, while false negatives occurred when relevant OUD information was documented in notes external to the health system (i.e., Epic’s “Care Everywhere”; see Discussion).

**Table 2.** Themes from Discordant Classifications.

|  | False Positive | False Negative |
| --- | --- | --- |
| Structured | - Urine toxicology resulted positive for fentanyl once without intentional use of opioid<br>- Chronic pain on opioid medication (including methadone or buprenorphine) | - Missed mention of overdose responsive to Narcan in Care Everywhere ED note<br>- History of “opiate withdrawal” in Care Everywhere<br>- Cardiac arrest related to “OUD” |
| LLM | - One suicide attempt by overdose of fentanyl without consistent use of opioids<br>- Opioid dependence in Problem List counted as OUD<br>- Counted withdrawal from kratom as opioid-like substance | - Missed mention of OUD on methadone as part of Care Everywhere<br>- Missed mention of overdose responsive to Narcan in Care Everywhere ED note<br>- Anesthesia Procedure Note mention of polysubstance abuse, including OUD<br>- OUD documented in Care Everywhere notes |
| Expert 1* | - Stimulant use mislabeled as OUD in provider notes<br>- Urine toxicology positive for fentanyl twice, but no explicit mention of intentional use of opioid | - Methadone abuse reported by the patient’s wife misclassified as methadone maintenance<br>- Missed mention of prior heroin addiction |
| Expert 2* | - Opioid dependence falsely categorized as OUD due to explicit mention of use disorder in provider notes | - Missed mention in provider note of regular opiate use combined with fentanyl positive on urine drug screen<br>- Patient on Butrans for chronic pain but multiple urine toxicology results with fentanyl positivity<br>- Missed mention of prior heroin addiction and opioid withdrawal |

**Abbreviations:** ED, emergency department; OUD, opioid use disorder; LLM, large language model
\*These represent discordant cases between Expert 1 and Expert 2

## LIMITATIONS

First, generalizability may be limited because the structured phenotype includes addiction consultation notes and expert reviewers accessed off-site EHR documentation through Care Everywhere, resources that may not be available across all health systems. This was intentional to strengthen the reference standard and reflect comprehensive clinical review. Second, as a tertiary care referral center, UCSF documentation practices may not reflect other hospital settings, potentially affecting the structured phenotype and LLM performance. Future research should validate these findings across different health systems and populations.

## DISCUSSION

Both the structured computable OUD phenotype and the LLM performed well for OUD identification compared to expert physician review. The structured phenotype demonstrated slightly higher sensitivity, whereas the LLM demonstrated near-perfect specificity and higher positive predictive value.

Given the low prevalence of OUD in our study population, which reflects real-world practice, predictive values are particularly relevant. Both approaches achieved high negative predictive value, supporting their ability to rule out OUD in most encounters. However, the LLM’s higher positive predictive value indicates fewer false positives, potentially reflecting greater ability to interpret clinical nuance within narrative documentation and reduce unnecessary alerts.

Prior studies have shown that LLMs can extract OUD-related concepts from clinical text and, in some settings, outperform earlier AI approaches for interpreting narrative documentation.^11–13^ However, emergency CDS tools have traditionally relied on rule-based computable phenotypes, built from diagnosis codes, medications, laboratory results, and keywords.^6,14^ Our study directly compares an LLM with such a structured phenotype. Although the structured phenotype demonstrated slightly higher sensitivity, the LLM achieved almost perfect specificity and a higher positive predictive value. These complementary performance characteristics suggest a staged CDS approach. A structured phenotype could serve as an initial step, followed by LLM review to improve precision and decrease false positives. In low-prevalence ED settings, this strategy may reduce alert fatigue while preserving case detection. Notably, this performance was achieved using documentation from a single ED visit, which is often not extensive. Thus, LLMs may be particularly valuable in busy clinical environments such as the ED, where time constraints limit comprehensive chart review and systematic human screening is difficult to sustain.

Future research should focus on refining reference standards and optimizing implementation across diverse clinical environments. Although expert agreement was high, disagreement in select cases underscores the clinical complexity of distinguishing OUD from physiologic dependence and other forms of opioid exposure. These diagnostic ambiguities affect both clinicians and automated systems; LLMs should not be held to a higher standard than human experts when classification itself is inherently challenging. Continued work on prompt engineering and more nuanced classification frameworks may improve model performance in clinically ambiguous scenarios. Prospective studies incorporating standardized diagnostic criteria and multidisciplinary adjudication may further strengthen outcome definitions. Future evaluations should also incorporate longitudinal records, including outside documentation available through health information exchange (e.g., Care Everywhere), and assess performance across multiple health systems and real-world workflows. Such efforts will help define how AI-assisted OUD identification can be responsibly integrated into ED-based clinical decision support.

## Data Availability

Given the sensitive nature of the data in the present study, it will not be made available.

