## Supplement for "Comparing computable structured phenotype- versus large language model-identification of opioid use disorder using electronic health record data"

**Appendix A. Large Language Model Opioid Use Disorder Identification Prompt Development**

Prompt development was conducted using a separate set of ten representative ED encounters selected to reflect a range of clinical scenarios related to opioid use and OUD. Initial prompt wording focused on identifying any mention of opioid-related substance use; early iterations resulted in false positives in cases involving prescribed opioids for acute pain or family history of OUD. The prompt was subsequently refined to clarify interpretive caveats (e.g., distinguishing prescribed opioid use for pain from misuse) and to require documentation meeting predefined clinical criteria for OUD. After refinement, the prompt was finalized and fixed prior to application to the full study cohort.

The model (OpenAI ChatGPT-4.1) was accessed via R (v4.4.0) using the OpenAI API. Each ED encounter was processed independently in a single request using deterministic inference (temperature = 0). No conversational memory was retained between encounters. The model was instructed to return a binary classification (YES/NO) and a brief rationale in a standardized format. Model outputs were stored verbatim and were not manually edited prior to analysis.

You are a clinical informatics specialist analyzing emergency department notes to see if this patient has documentation of opioid-use disorder (OUD) related conditions.

You will score all the notes provided as a whole set, and score as yes/no based on if any of the following criteria are documented in ANY the note(s):

1. history of opioid use disorder (OUD) or current OUD,
2. Current or past illicit opioid use (e.g., heroin, fentanyl, non-prescribed opioids),
3. Opioid overdose (unless clearly accidental use of pain medications),
4. Medication for opioid use disorder unless stated that it is specifically prescribed for pain (e.g. buprenorphine, methadone, naltrexone),
5. Documented history of IV drug use (IVDU) involving opioids or any IVDU when the drug not otherwise specified.

Specific caveats: Do not count as opioid use disorder if current or prior opioids/opiates were prescribed for pain (including methadone for pain), unless stated the opiate was abused, and do not count substance abuse limited to non-opioid substances (methamphetamine, cocaine, marijuana).

The patient's ED notes are here: "…”

'The response should be in this format (1 response per prompt):

Yes, 1 or more of the criteria were met anywhere in any of these notes.

No, none of the criteria were met.

Provide your reasoning here: “….”

**Appendix B. Patient Demographic Characteristics of ED Encounters**

|  | **Total Encounters**  **N (%)** | **OUD Encounters**  **n (%)** | **Non-OUD Encounters**  **n (%)** |
| --- | --- | --- | --- |
| **Overall** | 506 (100%) | 303 (60%) | 203 (40%) |
| **Age, Mean (Std. Dev.)** | 50.6 (18.2) | 49 (16.5) | 53.1 (20.2) |
| **Race and Ethnicity** |  |  |  |
| Asian | 71 (14%) | 20 (6.6%) | 51 (25.1%) |
| Black | 92 (18.2%) | 56 (18.5%) | 36 (17.7%) |
| Latinx | 69 (13.6%) | 37 (12.2%) | 32 (15.8%) |
| Multi-racial | 21 (4.2%) | 15 (5%) | 6 (3%) |
| Other^a^ | 29 (5.7%) | 25 (8.3%) | 4 (2%) |
| White | 223 (44.1%) | 149 (49.2%) | 74 (36.5%) |
| **Sex** |  |  |  |
| Male | 308 (60.9%) | 196 (64.7%) | 112 (55.2%) |
| Female | 190 (37.5%) | 99 (32.7%) | 91 (44.8%) |
| Other^b^ | 8 (1.6%) | 8 (2.6%) | 0 (0%) |
| **Insurance** |  |  |  |
| Medicaid | 164 (32.4%) | 142 (46.9%) | 22 (10.8%) |
| Medicare | 169 (33.4%) | 96 (31.7%) | 73 (36%) |
| Commercial | 135 (26.7%) | 43 (14.2%) | 92 (45.3%) |
| Other^c^ | 38 (7.5%) | 22 (7.3%) | 16 (7.9%) |
| **Language: Interpreter Needed?** | 34 (6.7%) | 11 (3.6%) | 23 (11.3%) |

**Abbreviations:** OUD, opioid use disorder

^a^Included “Other”, “Unknown/Declined”, “Southwest Asian and North African”

^b^Included “Choose not to disclose”, “Declines to answer”, “Something else”, “Unknown”

^c^Included “Other” and “Worker’s Comp”
